# Brain injury dating via histological examination of vascular changes in parenchymal tissue: A pilot study

**DOI:** 10.1101/2025.03.26.25324626

**Authors:** Ashok k Rastogi, Dinil D Nangelil, Rajesh Kumar, Tarun Kumar, Sanjeev K Paikra, Toshal D Wandhade

**Author notes:** Corresponding author Department of Forensic Medicine & Toxicology, All India Institute of Medical Science, Patna (Bihar), India, Email ID.

## Abstract

**Background:** Traumatic brain injury (TBI) is a complex multidisciplinary medical, biological, and social issue due to its persistently high mortality and disability rates, particularly among individuals of working age. Determining the survival interval following a TBI is often of medicolegal significance, requiring targeted tissue sampling and histological evaluation. Neuronal and vascular changes observed in injured brain tissue can provide valuable insights into the timing of the injury.

**Aims & Objectives:** To estimate the timing of traumatic brain injury based on vascular changes observed through histological examination.

**Methods:** This study included cases of TBI with varying survival intervals alongside non-injured controls. Macroscopic examination of brain specimens was performed during autopsy, followed by histological analysis of harvested brain tissue. Tissue sections were stained with Hematoxylin and Eosin and examined under a light microscope.

**Conclusion:** Histological analysis is a valuable tool for complementing macroscopic diagnoses, especially when gross evidence of intracranial trauma is absent. It can also distinguish traumatic from non-traumatic lesions and assist in estimating the survival interval after a head injury. This information can be crucial in medicolegal investigations, as the biological time of death may not always align with the legal time of death.

## Introduction

Traumatic brain injury (TBI) is a significant medical challenge, particularly among working-age individuals, due to its high rates of mortality and disability. (1,2) Determining the survival interval after a TBI requires targeted tissue sampling and meticulous histological evaluation. (3) Histological examination can visualize neuronal and vascular changes, providing crucial insights into the timing of injury. Following a TBI, inflammation occurs rapidly in response to brain tissue damage, regardless of the injury’s mechanism or severity. (4) Sequential inflammatory changes can help estimate the time elapsed since the injury. Additionally, TBI disrupts the blood-brain barrier (BBB), leading to altered permeability. (5)The BBB consists of three key cell types: endothelial cells, pericytes, and astrocytes. TBI can result from direct insults, often accompanied by microvascular changes. These alterations may be subtle and present even in areas without visible traumatic injury or disrupted tight junctions. (6) Insults to the brain can trigger the release of autocrine and paracrine factors, compromising vascular integrity. (7) Vascular dysfunction has been implicated in second-impact syndrome following a TBI. Additionally, many TBI patients experience a significant reduction in cerebral blood flow (CBF), further contributing to neurological impairment. (8)

## Aim and objective

Histological Dating of Traumatic Brain Injury Based on Vascular Changes.

## Methodology

This study included 12 traumatic brain injury (TBI) cases with varying survival durations and four non-injured control cases. Macroscopic examination of brain specimens was performed during autopsy, followed by histological analysis of harvested brain tissue. Tissue sections were stained with Hematoxylin and Eosin and examined under a bright-field light microscope. Ethical clearance was obtained and informed written consent was secured before sample collection.

## Observations

We analyzed 12 cases of traumatic brain injury (TBI) resulting from road traffic accidents, with varying survival durations, along with four cases of non-traumatic brain tissue as controls. In the control samples, taken from deceased individuals with varying postmortem intervals, the only notable observations were congestion and vacuolation in and around the vessels. No other significant histological findings were observed.

**Table 1:** Details of the test samples with respect to time since death and injury duration.

| SN | Time since death | Time since injury |
| --- | --- | --- |
| 1. | 04hr | 12 hr |
| 2. | 09hrs | 12hrs |
| 3. | 03hrs | 12hrs |
| 4. | 18hrs | 08D |
| 5. | 06hrs | 03D |
| 6. | 12hrs | 06hrs |
| 7. | 12hrs | 06D |
| 8. | 12hrs | 01D |
| 9. | 12hrs | 03D |
| 10. | 06hrs | 04hrs |
| 11. | 12hrs | 04D |
| 12. | 12hrs | 4hrs |

**Table 2:** Time since death in Control cases.

| SN | Cause of death | TSD |
| --- | --- | --- |
| 1. | Hanging | 12hrs |
| 2. | Electrocution | 17hrs |
| 3. | Electrocution | 20hrs |
| 4. | Hanging | 12hrs |

**Table 3:** Observation of histology in brain tissue at varying time intervals after injury.

| SN | Feature | Control sample | TSI 0-06 hours | TSI 06-12 hours | TSI 12-24 hours | TSI 1-3 days | TSI 3-6 days | TSI 08 days |
| --- | --- | --- | --- | --- | --- | --- | --- | --- |
|  | No cases | 04 | 03 | 03 | 01 | 02 | 02 | 01 |
| 1) | Vessel congestion | **P4** | P | P | P | P | P | P |
| 2) | Perivascular edema | --- |  | **P1** | -- | **P1** |  | **P1** |
| 3) | Perivascular Hemorrhagic | --- | **P2** | **P3** | P1 | P1 |  |  |
| 4) | Endothelial oedema | --- | P2 | **P3** | P1 | P2 | -- | -- |
| 5) | Vacuoles in the vessels | 03 | ---- | -- | --- | P2 | P1 |  |
| 6) | Neutrophil infiltration | --- | --- | --- | P1 |  |  |  |
| 7) | Macrophage infiltration(microglia) | --- | P1 | P2 | -- | **P1** | **P1** |  |
| 8) | Lymphocyte infiltration | --- | --- | -- | --- | P1 | P1 |  |

**Figure 1:**
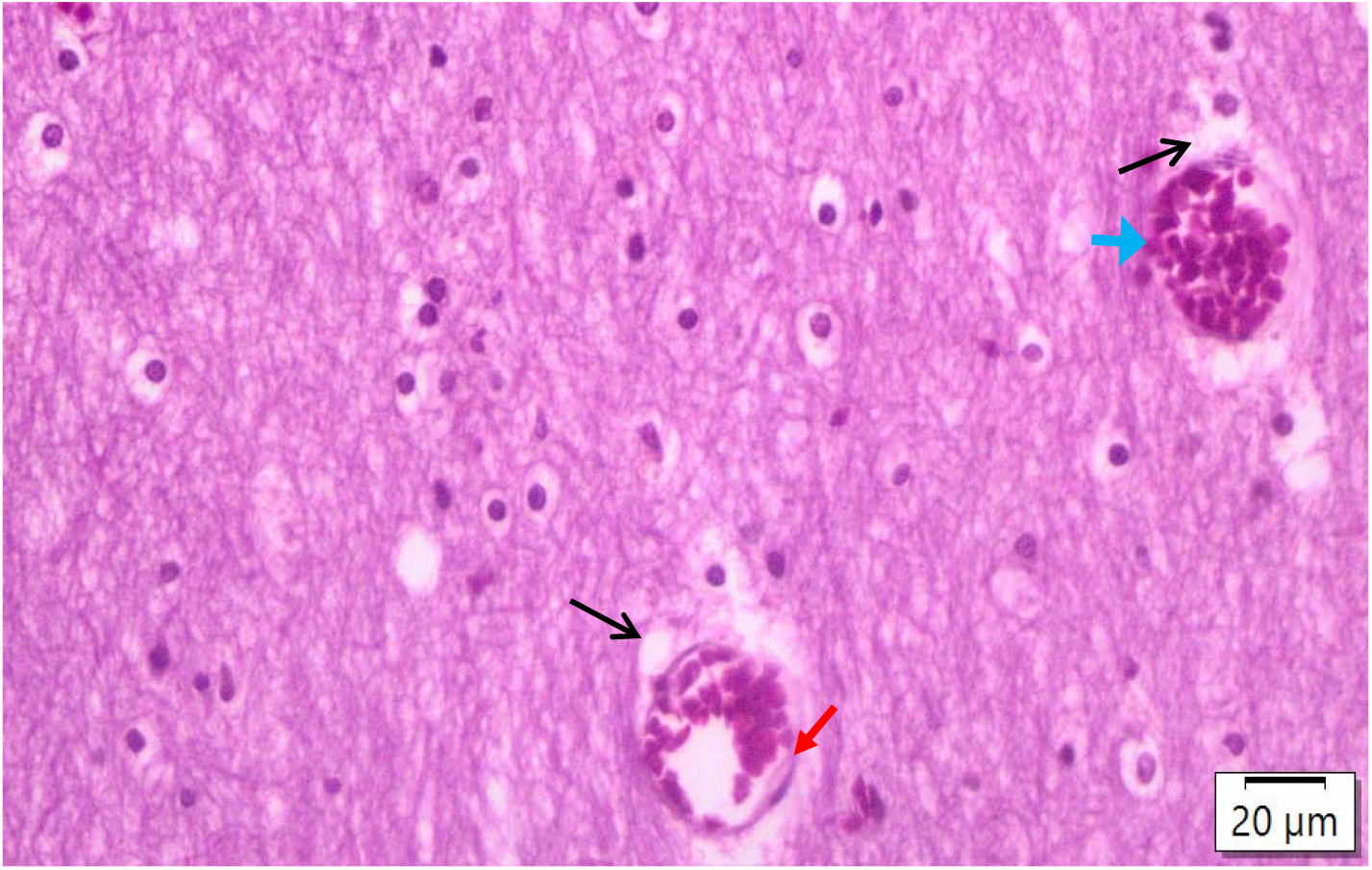
Congestion of the blood vessels (Blue arrow), perivascular vocalisations (black arrow) and intravascular vacuolation (Red arrow) in control cases of death due to electrocution cases. The TSD is 20 hours.

**Figure 2:**
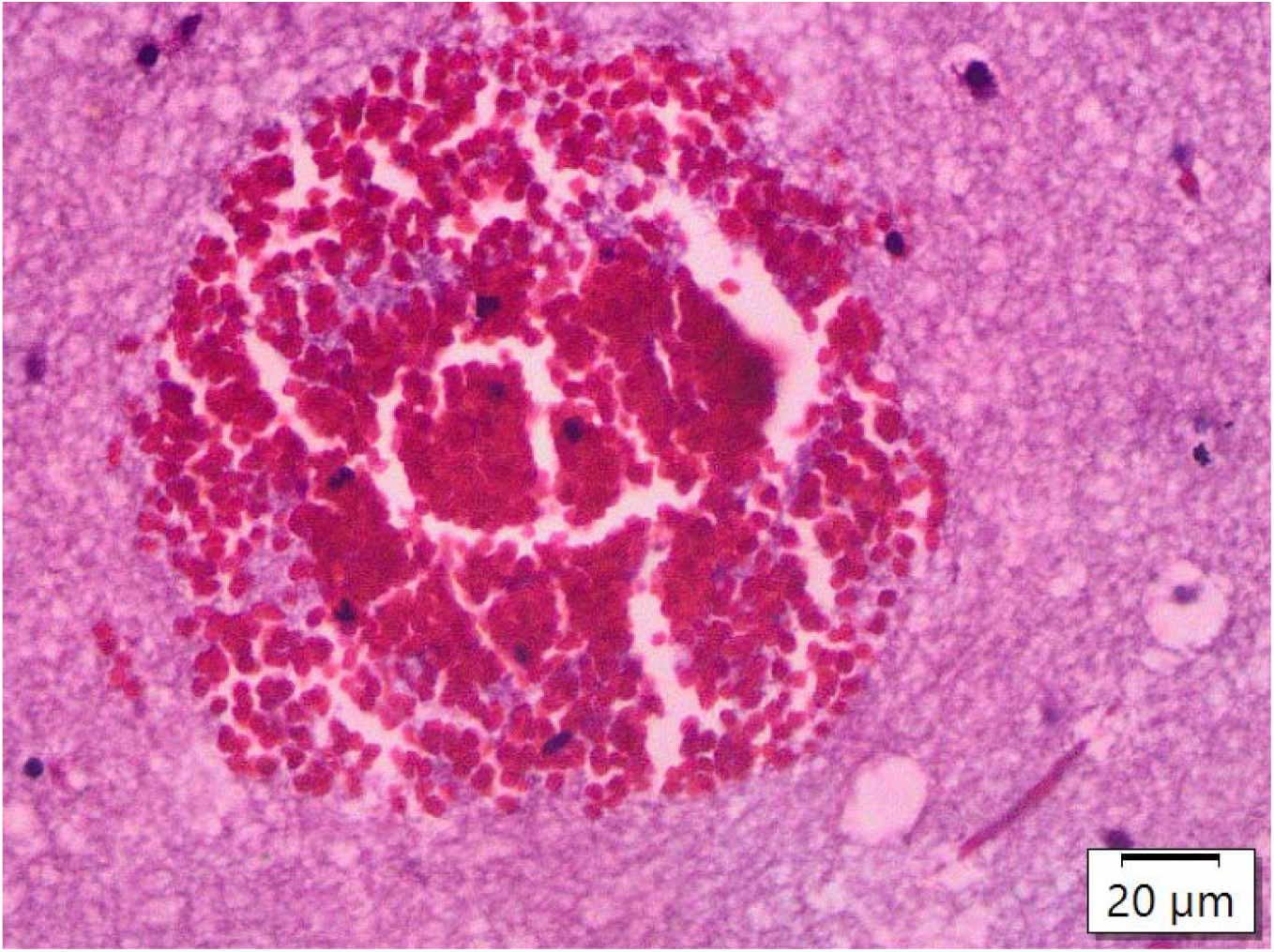
Blood collection in the parenchyma TSI 12 hours and TSD 03 hours.

**Figure 3:**
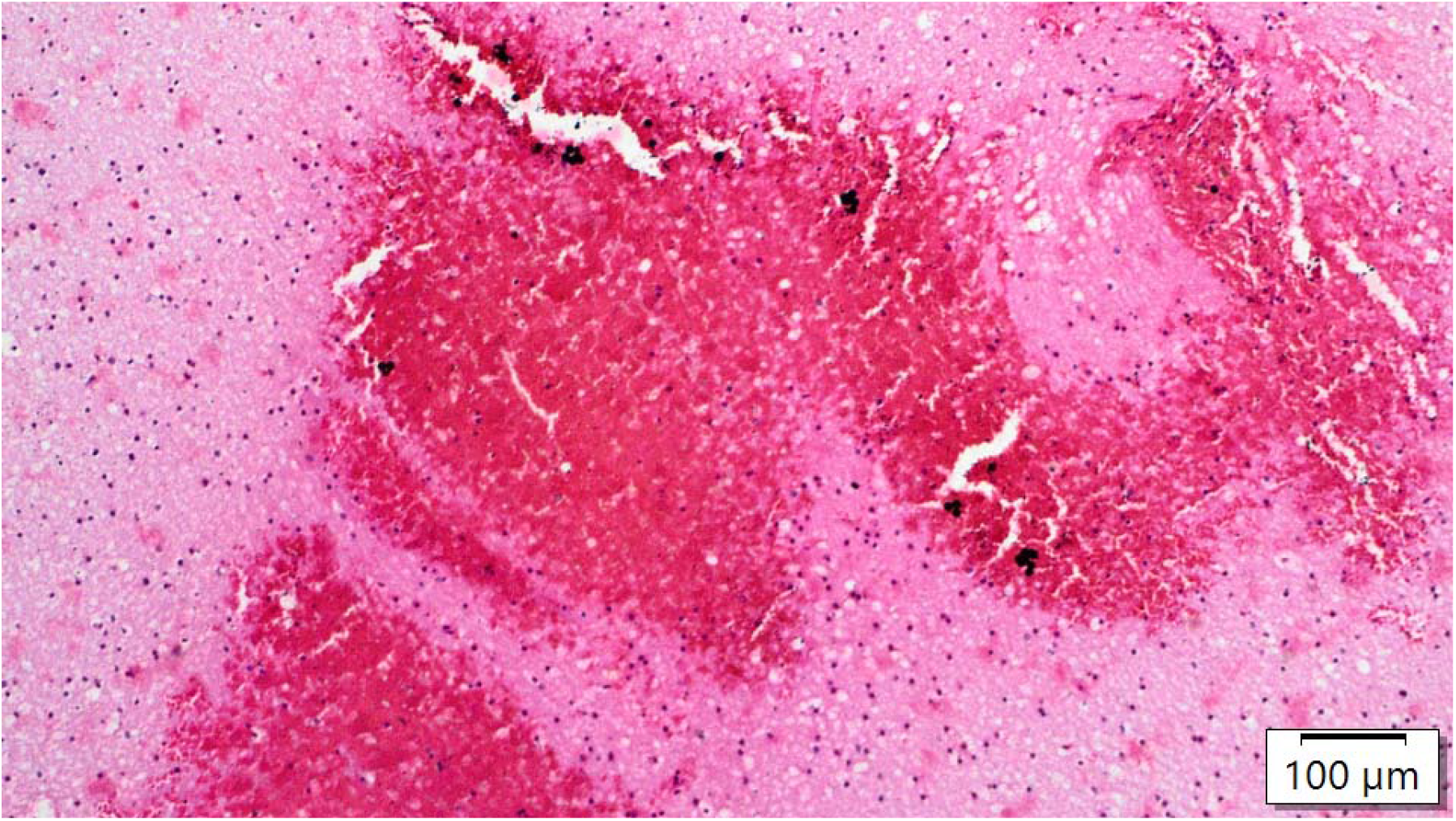
Multiple varying sizes of parenchymal haemorrhage, TSI 04 hours and TSD 12 hours.

**Figure 4:**
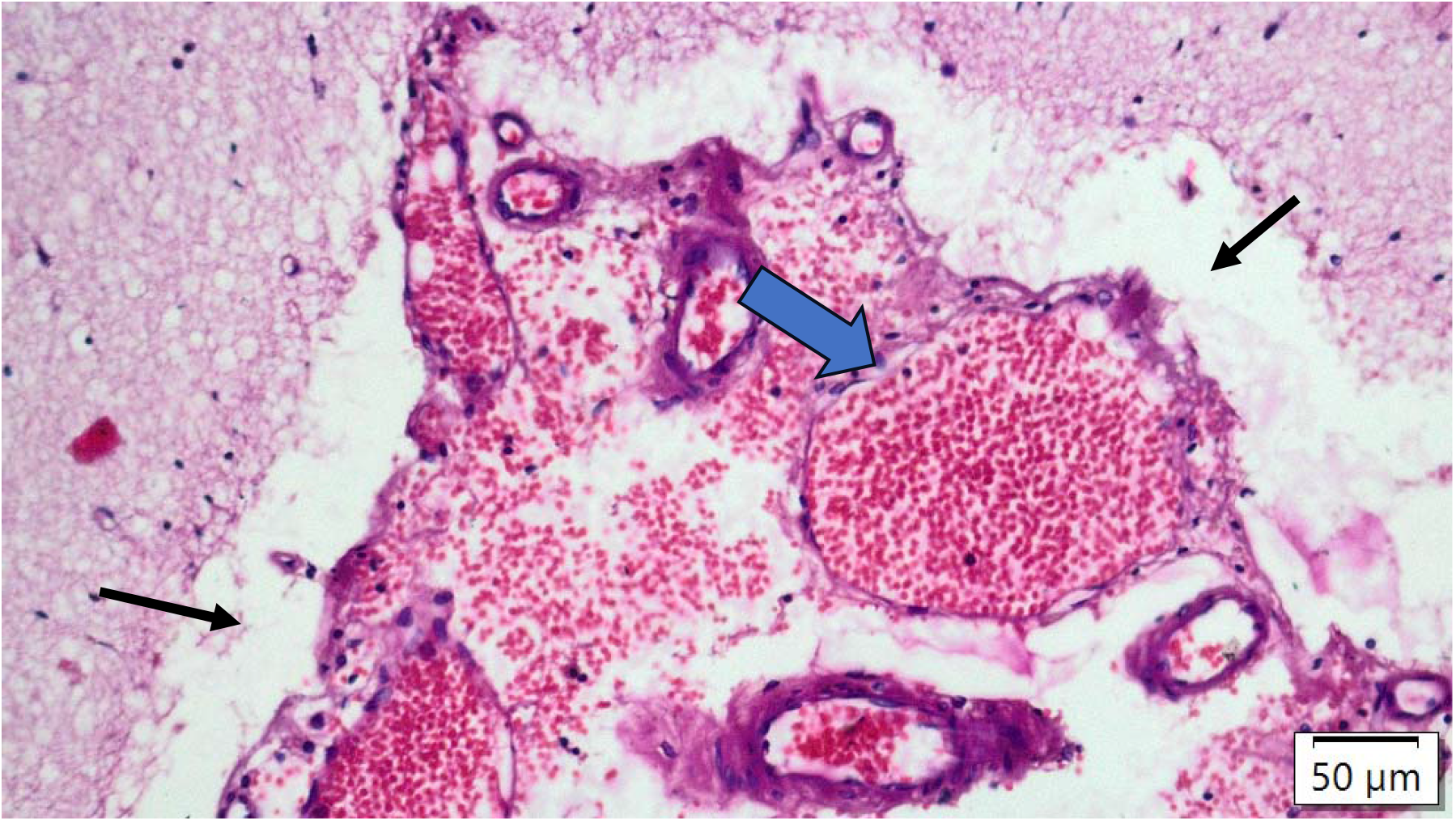
Perivascular vacuolation (black arrow), vacuolation inside the vessel (Blue arrow), TSI 12 hours and TSD 09 hours

**Figure 5:**
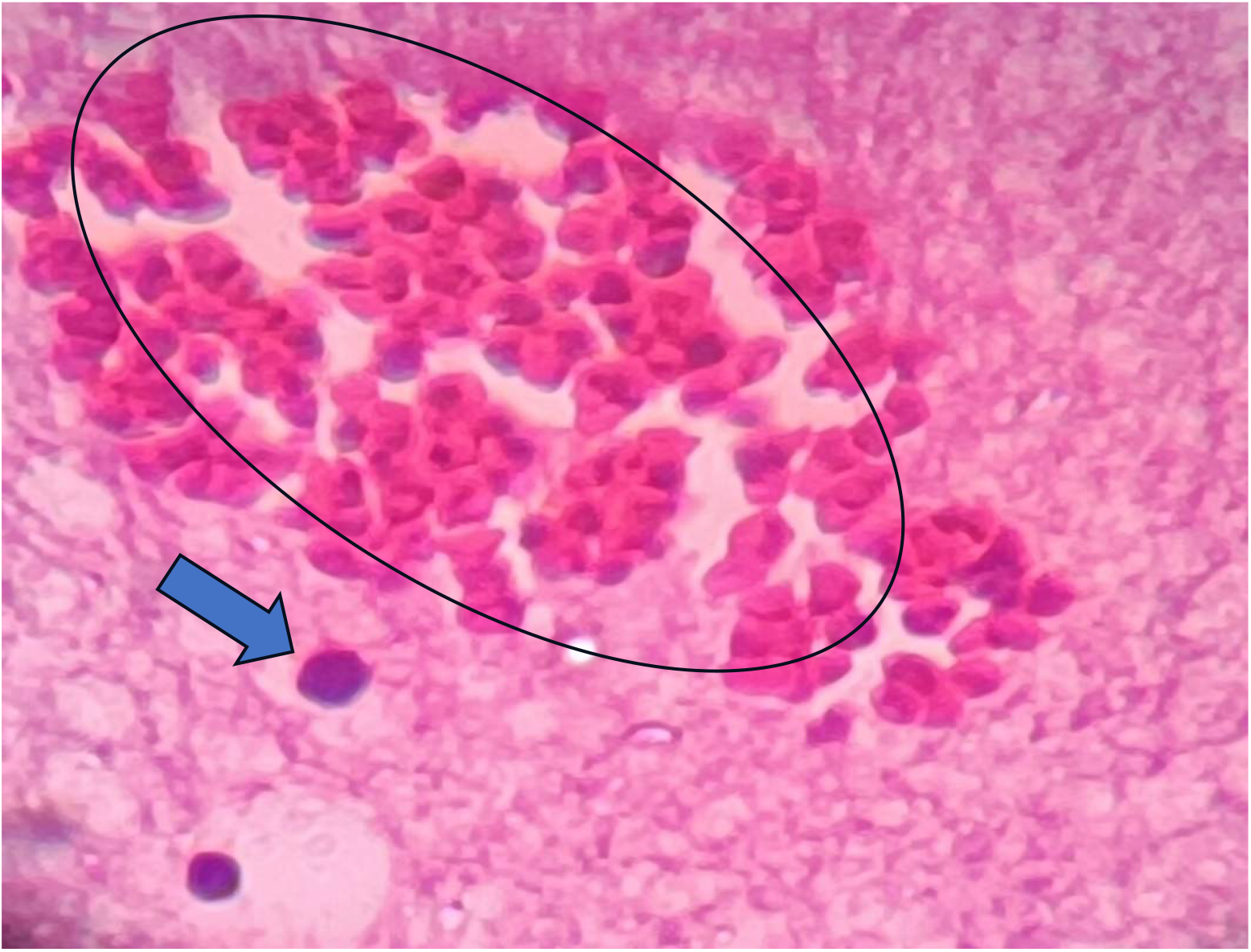
Collection of blood in parenchymal tissue (black circle) and lymphocyte infiltration(Blue Arrow), magnification 400X.

**Figure 6:**
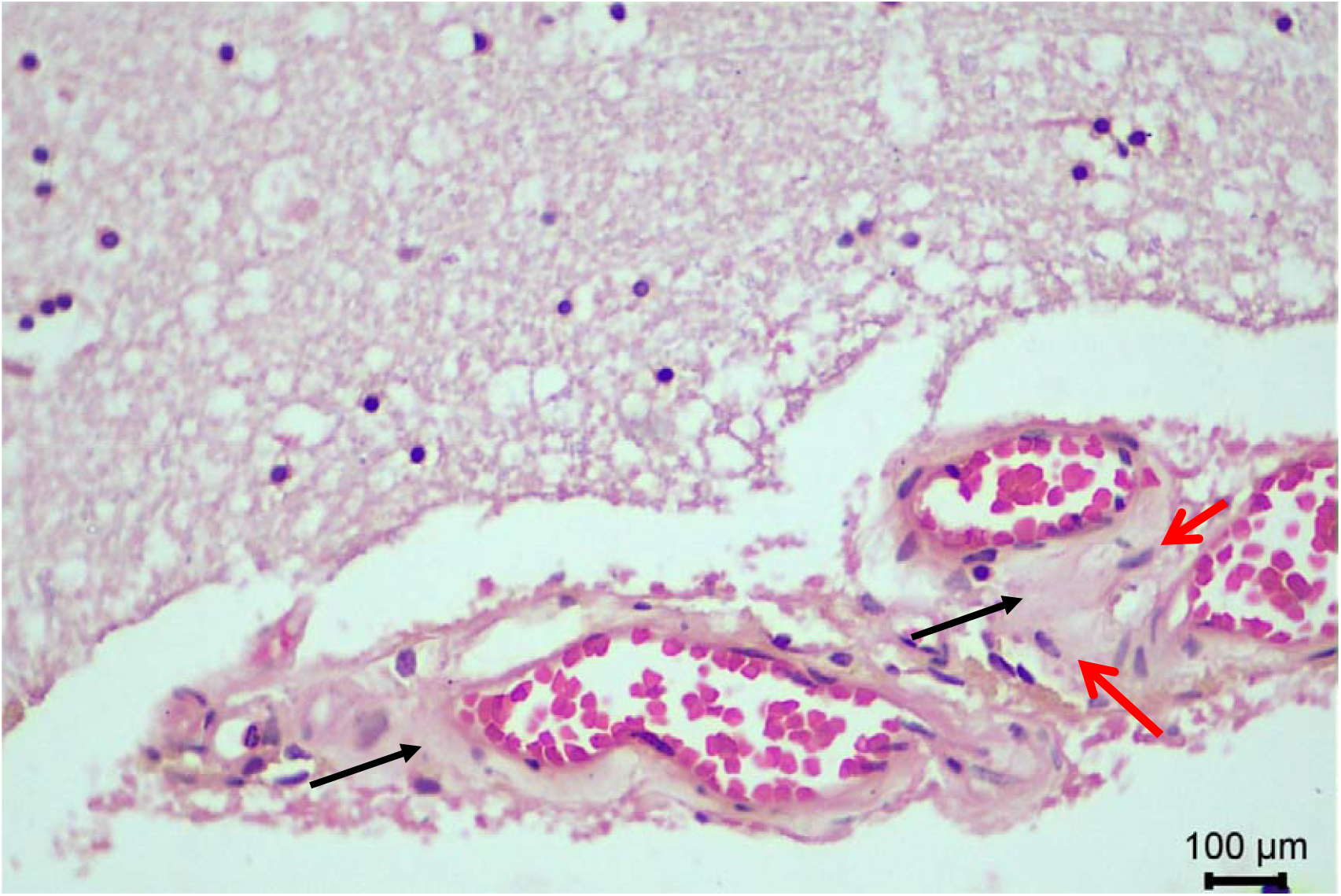
Perivascular oedema (Black Arrow), macrophage infiltration in the form of microglia around the vessels (Red arrow), TSI: 3 days, TSD: 12 hours.

**Figure 7:**
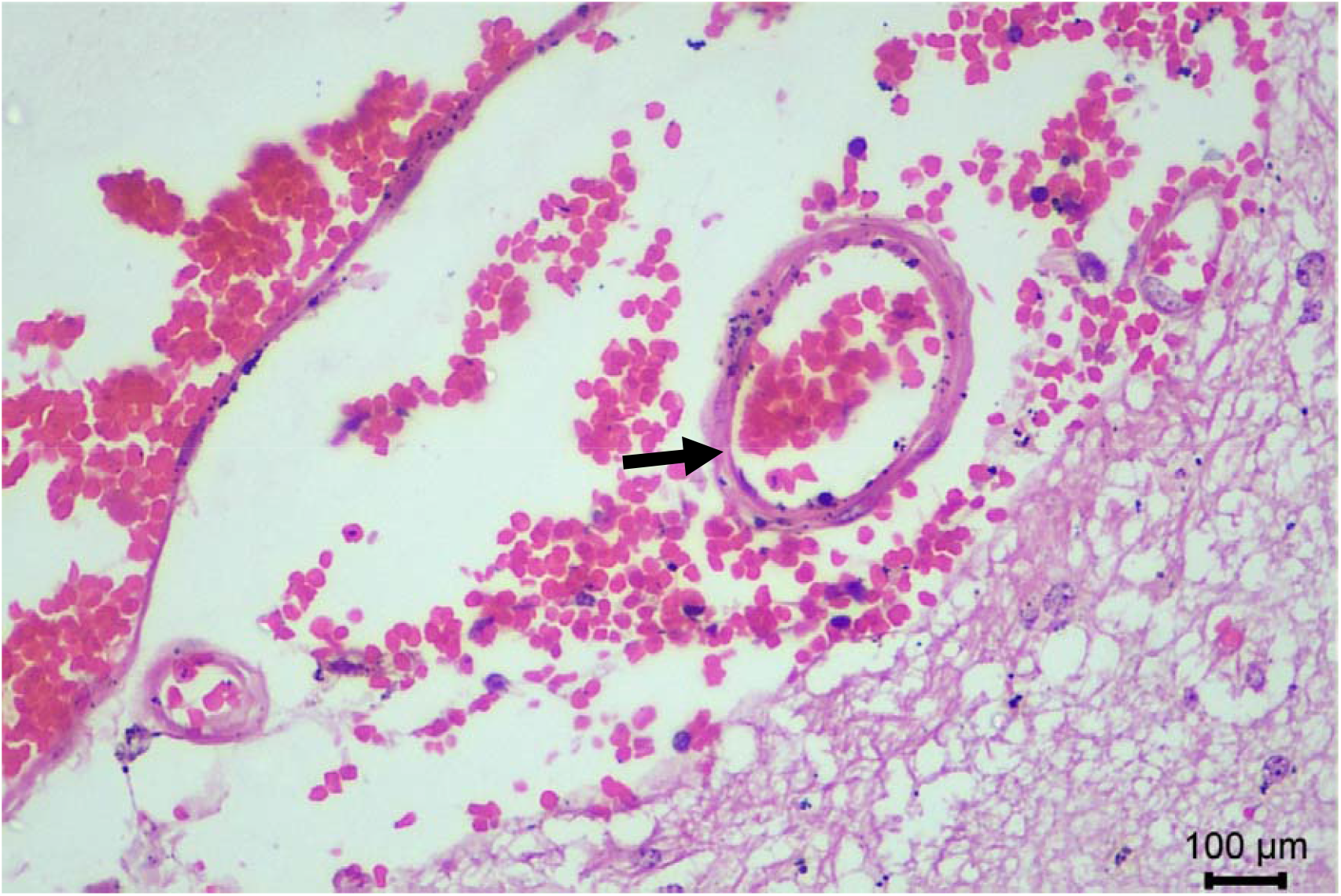
Endothelial oedema (Black Arrow), TSD-12 hours, TSI-04 days

**Figure 8:**
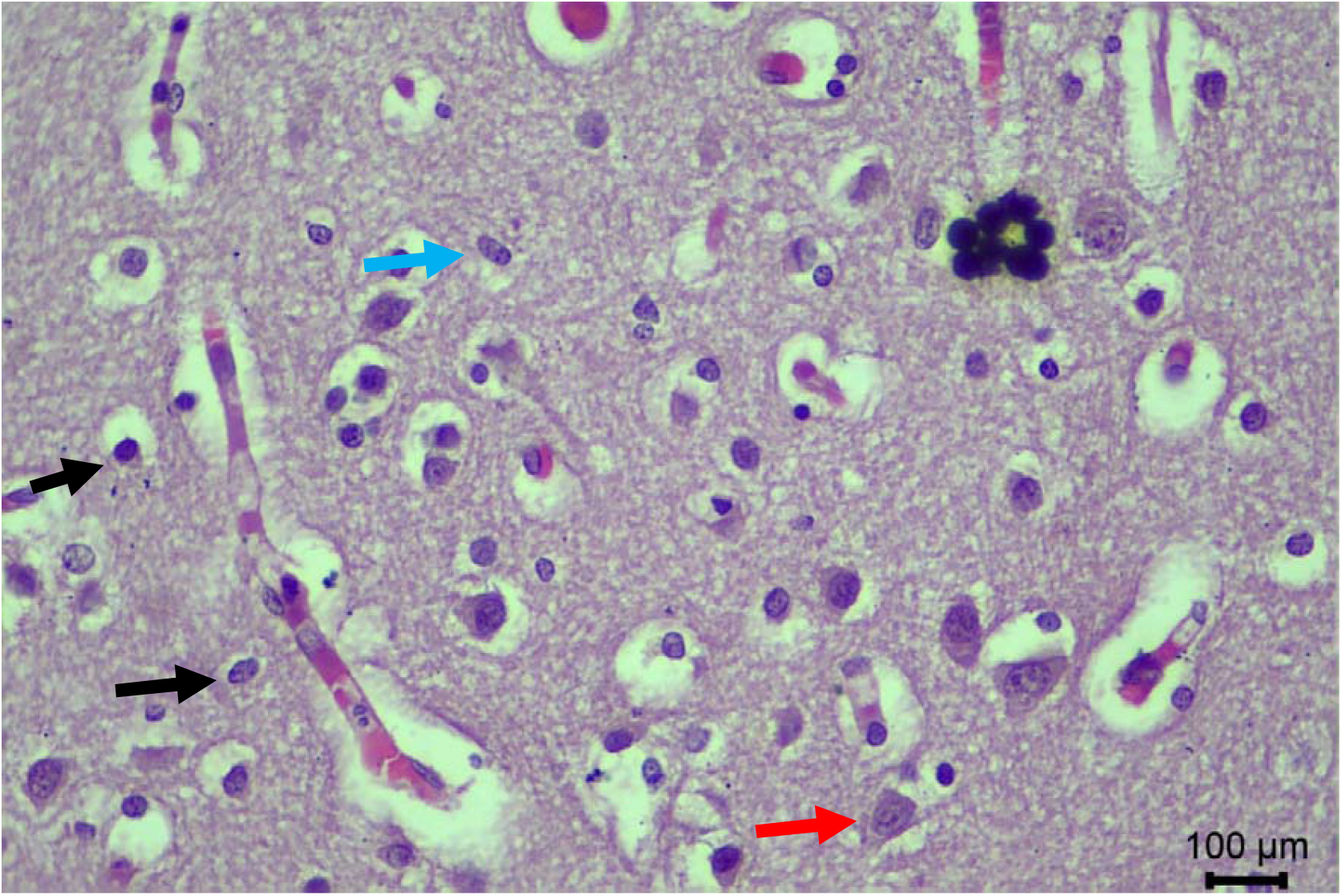
Brain parenchyma showing lymphocytic infiltrate (Black Arrow) along with reactive glial cells (Red Arrow) & microglia (Blue Arrow), TSD-12 hours, TSI-04 days

### Theoretical Importance

The healing of brain injuries involves a process that includes inflammation, collagen formation, and neovascularisation over time.

### Data Collection and Analysis Procedures

After staining the injured brain tissue with hematoxylin and eosin, changes and findings were noted and analyzed under a light microscope to determine the time since the head injury.

### Question Addressed

How do we estimate the time since a traumatic brain injury caused by physical violence or a road traffic accident (RTA)?

### Ethical Statement

Ethical clearance was obtained wide letter, No. AIIMS/Pat/IEC/2024/2, from the Institutional Ethics Committee. Before collecting the sample, informed written consent was taken from the relatives of the deceased.

## Discussion

It is essential to recognise that Traumatic Brain Injury (TBI) is a process rather than a one-time event. (9) Trauma can damage the blood-brain barrier, causing secondary injury and peripheral exposure to the brain. (10) Neither axonal nor vascular changes were found with a 10-hour interval between injuries. (11) Brain injuries can result in vascular dysfunction, which can lead to both morbidity and mortality. (12)

If a person who has suffered a severe head injury dies within minutes, there may be widespread petechial haemorrhages throughout the brain as diffuse vascular injury. (13) After traumatic brain injury in rats, early structural changes were observed at the endothelium. Electron microscopy of the evolving damage revealed perivascular oedema that resolved within 48 hours. (6) Identifiable pericytes were found to migrate from the vessel into the parenchyma. It was observed that pericytes, which can be identified, migrated from the blood vessel into the surrounding parenchymal tissue. (6) In some blood vessels after TBI, the blood vessel is enlarged and filled with blood, causing congestion, while the endothelial cells that line the vessel are swollen. (14) Polymorphonuclear cells were prominent and easily noticeable in some blood vessels. Neutrophils can be observed leaving blood vessels and entering damaged tissue within hours of injury. During the early post-injury period, both intravascular neutrophil margination and extravasation can occur. Leukocytes remain motile for up to 6-8 hours post-mortem. (22) From 4 to 16 hours, the neutrophilic reaction intensifies, peaking at about three days,(25) With a survival time of 1 to 4 hours, a few infiltrating neutrophils appear, and erythrocytes begin to lyse. (26)

Martin et al. (1997) found that severe head injury causes hypoperfusion in the first 24 hours, followed by hyperemia during days 1-3, and vasospasm accompanied by hypoperfusion on days 4-15.(15) After a brain injury, vessel endothelial cells develop vacuoles and pinocytotic vesicles. (16) In rabbits, Leukocyte accumulation and abnormal vasodilation were detected within 30 minutes of fluid percussion TBI.(17) A contusion is commonly recognised as the defining characteristic of a head injury. (18,19) Damage to blood vessels and surrounding tissue, including cortical and sub-cortical tissue, is characterised by streaks or clusters of small haemorrhages in areas of superficial damage.(20) Intracranial haemorrhages often occur due to the rupture of intrinsic cerebral vessels, resulting in cerebral contusions.(21) Hemorrhage begins immediately at perivascular sites, extends, and typically resolves within five days, though larger extravasations may persist for weeks to months. Oedema develops within minutes, peaks over several hours, stabilizes for a few days and subsides by day six. Lymphocytes appear around days 3-4 and may persist. Hemosiderin-laden macrophages emerge in small numbers by day five but become more apparent after seven days. (23)

Phagocytosis occurs between 12 and 24 hours after injury but is not prominent. Red neurons may persist at the contusion periphery for up to 6 months.(23) The first appearance of macrophages containing hemosiderin occurs after 6-7 days of survival. (24) Capillary proliferation seen, which commences at about 5 to 7 days postinjury. (23) Delayed traumatic brain injury (TBI) occurs as a complication of primary brain damage, exacerbating altered homeostasis over hours, days, or weeks. (23) After seven days, the number of neutrophils and lymphocytes/macrophages is similar. Intact erythrocytes are present but are lysed by days 9 to 20. (26,27)

## Data Availability

All data produced in the present work are contained in the manuscript

## Limitation of the study

A stratified sample size would be considered for more accurate results and conclusions.

## Conclusion

Forensic pathologists rely on gross findings for TBI diagnosis, but histology provides crucial evidence, especially when trauma is not visibly apparent. It aids in distinguishing non-traumatic lesions, estimating survival intervals, and clarifying discrepancies between biological and legal time of death, which are critical factors in legal proceedings.

## Acknowledgement

The authors are grateful to the family of the deceased, who provided consent for the collection and reporting of clinical data.

## Funding agency

None

## Authors’ contributions

AKR and RK conceptualised the study. AKR, TDW and DDN carried out data collection. AKR, DN and SKP performed laboratory work. AKR, TK, and RK conducted pathological validation and analysed the data. AKR and RK wrote the first draft of the manuscript. All authors reviewed and approved the final version of the manuscript.

